# Improving reporting standards for polygenic scores in risk prediction studies

**DOI:** 10.1101/2020.04.23.20077099

**Authors:** Hannah Wand, Samuel A. Lambert, Cecelia Tamburro, Michael A. Iacocca, Jack W. O’Sullivan, Catherine Sillari, Iftikhar J. Kullo, Robb Rowley, Jacqueline S. Dron, Deanna Brockman, Eric Venner, Mark I. McCarthy, Antonis C. Antoniou, Douglas F. Easton, Robert A. Hegele, Amit V. Khera, Nilanjan Chatterjee, Charles Kooperberg, Karen Edwards, Katherine Vlessis, Kim Kinnear, John N. Danesh, Helen Parkinson, Erin M. Ramos, Megan C. Roberts, Kelly E. Ormond, Muin J. Khoury, A. Cecile J.W. Janssens, Katrina A.B. Goddard, Peter Kraft, Jaqueline A. L. MacArthur, Michael Inouye, Genevieve Wojcik

## Abstract

Polygenic risk scores (PRS), often aggregating the results from genome-wide association studies, can bridge the gap between the initial discovery efforts and clinical applications for disease risk estimation. However, there is remarkable heterogeneity in the reporting of these risk scores. This lack of adherence to reporting standards hinders the translation of PRS into clinical care. The ClinGen Complex Disease Working Group, in a collaboration with the Polygenic Score (PGS) Catalog, have updated the Genetic Risk Prediction (GRIPS) Reporting Statement to the current state of the field and to enable downstream utility. Drawing upon experts in epidemiology, statistics, disease-specific applications, implementation, and policy, this 22-item reporting framework defines the minimal information needed to interpret and evaluate a PRS, especially with respect to any downstream clinical applications. Items span detailed descriptions of the study population (recruitment method, key demographic and clinical characteristics, inclusion/exclusion criteria, and outcome definition), statistical methods for both PRS development and validation, and considerations for potential limitations of the published risk score and downstream clinical utility. Additionally, emphasis has been placed on data availability and transparency to facilitate reproducibility and benchmarking against other PRS, such as deposition in the publicly available PGS Catalog. By providing these criteria in a structured format that builds upon existing standards and ontologies, the use of this framework in publishing PRS will facilitate translation of PRS into clinical care and progress towards defining best practices.

**Summary:** In recent years, polygenic risk scores (PRS) have increasingly been used to capture the genome-wide liability underlying many human traits and diseases, hoping to better inform an individual’s genetic risk. However, a lack of adherence to existing reporting standards has hindered the translation of this important tool into clinical and public health practice; in particular, details necessary for benchmarking and reproducibility are underreported. To address this gap, the ClinGen Complex Disease Working Group and Polygenic Score (PGS) Catalog have updated the Genetic Risk Prediction (GRIPS) Reporting Statement into the 22-item Polygenic Risk Score Reporting Statement (PRS-RS). This framework provides the minimal information expected of authors to promote the validity, transparency, and reproducibility of PRS by encouraging authors to detail the study population, statistical methods, and potential clinical utility of a published score. The widespread adoption of this framework will encourage rigorous methodological consideration and facilitate benchmarking to ensure high quality scores are translated into the clinic.

## Main Text

The predisposition to common diseases and traits arises from a complex interaction between genetic and nongenetic factors. In the past decade there has been enormous success at discovering the landscape of disease-associated genetic variants as a result of the many collaborative consortia and large cohorts of well-phenotyped individuals and matched genetic information.^1–5^ In particular, genome-wide association studies (GWAS) have emerged as a powerful approach to identify disease- or trait-associated genetic variants and yielded summary statistics that describe the magnitude (effect size) and statistical significance of association between an allele and the outcome of interest.^4,6^ GWAS have been applied to a wide range of complex human traits and diseases, including height, blood pressure, cardiovascular disease, cancer, obesity, and Alzheimer’s disease.

The associations identified via GWAS can be combined to quantify genetic predisposition to a heritable trait, and this information can be used to conduct disease risk stratification or predict prognostic outcomes and response to therapy.^7,8^ Typically, information across many variants is used to form a weighted sum of allele counts, where the weights reflect the relative magnitude of association between variant alleles and the trait or disease. These weighted sums can include up to millions of variants, and are frequently referred to as **polygenic risk score(s) (PRS)**, or **genetic or genomic risk score(s) (GRS)**, if they refer to risk estimates of disease outcomes; or, more generally, **polygenic score(s) (PGS)** when referring to any outcome (*see* ***Box 1*** *for further discussion of nomenclature*). While there is active development of algorithms to decide how many and which variants to include and how to weigh them so as to maximize the proportion of variance explained or disease discrimination, there is an emerging consensus that the inclusion of variants beyond those meeting stringent GWAS significance levels can boost predictive performance.^9,10^ Methodological research has also established theoretical limits of PGS/PRS performance based on the trait’s genetic architecture and heritability.^11–15^

In the last decade, the landscape of genetic prediction studies has transformed. There have been over 900 publications mentioning PGS/PRS with significant developments in how PGS are constructed and evaluated, as well as many new proposed uses. The data available in the current era of biomedical research is larger and more consolidated than ever before. Biobanks and large-scale consortia have become dominant, yet frequently researchers have limited access to individual-level data. Since individual data is unavailable, most PRS risk predictions are developed from summary-level data (e.g. GWAS summary statistics) in secondary datasets, each of which come with their own specific methodological considerations.^16–18^ At the same time, there has been a push towards open data sharing as outlined in the FAIR (Findable, Accessible, Interoperable and Reusable) Data Principles ^3,19^, with an emphasis on ensuring that research is reproducible by all.

### Box 1.

**Definitions of relevant genetic risk prediction terms**

#### Polygenic Score(s) (PGS)

a single value that quantifies an individual’s genetic predisposition to a trait. Typically calculated by summing the number of trait-associated alleles in an individual weighted by per-allele effect sizes from a discovery GWAS, and normalized using a relevant population distribution. Sometimes referred to as a genetic score.

#### Polygenic Risk Score(s) (PRS)

a subset of PGS which is used to estimate risk of disease or other clinically relevant outcomes (binary or discrete). Sometimes referred to as a genetic or genomic risk score (GRS). See categories of PRS below.

#### Integrated Risk Model

a risk model combining PRS with other established risk factors for the outcome of interest, such as demographics (often age and sex), anthropometrics, biomarkers, and clinical measurements to estimate a specific disease risk.

#### Categories of use for PRS and/or integrated risk models

The addition of PRS to existing risk models has several potential applications, summarized below. In each, the aim of PRS integration is to improve individual or subgroup classification to the extent that there is meaningful clinical benefit.

**Disease Risk Prediction** – used to estimate an individual’s risk of developing a disease, based on the presence of certain genetic and/or clinical variables.

**Disease Diagnosis** – used to classify whether an individual has a disease, or a disease subtype, linked to a certain etiology based on the presence of certain genetic and/or clinical variables.^9,20^

**Disease Prognosis** – used to estimate the risk of further adverse outcome(s) *subsequent to* diagnosis of disease.^21^

**Therapeutic** – used to predict a patient or subgroup’s response to a particular treatment.^22^

The capacity of PRS to quantify genetic predisposition for many clinically relevant traits and diseases has begun to be established, with multiple potential clinical uses in settings related to disease risk stratification as well as proposed prognostic uses (e.g., predicting response to intervention/treatment). The readiness of PRS for implementation varies among outcomes, with only a few diseases, like coronary heart disease (CHD) and breast cancer, having mature PRS with potential clinical utility (**Boxes 2** and **3**, respectively). There has also been a rapid rise of direct-to-consumer assays and for-profit companies (23andMe, Color, MyHeritage, etc.) providing PGS/PRS results to customers outside of the traditional patient-provider framework. These concomitant advances have resulted in healthcare systems developing new infrastructures to deliver genetic risk information.^23^ These advances, individually and combined, have raised significant challenges for PRS reporting standards; from the very basic (e.g. reporting performance metrics on an external validation dataset) to the complicated (e.g. making the raw variant and weight information for a PRS available), necessitating the updating of existing standard for reporting genetic risk prediction studies to convey the increased scope of PRS and complexity for their clinical applications.

### Box 2

**Current CHD PRS and their potential uses**

Many PRS have been developed for CHD, varying in the computational methods used, the number of variants included (50–6,000,000), and the GWAS and cohorts used for PRS training. For example, the latest and currently most predictive CHD PRS use GWAS summary statistics from the CardiogramPlusC4D study ^24^, and mainly differ by the computational methods used to select and weight individual variants (including LDpred ^25,26^, lassosum ^27^, and meta-scoring approaches ^28^, and how they are combined into risk models with conventional risk factors. These PRS may provide useful information for predicting risk of CHD that is largely orthogonal to conventional risk factors (age, sex, hypertension, cholesterol, BMI, diabetes, smoking) as well as family history. Clinical applications may include:

- Improved prediction of risk for future adverse cardiovascular events when added to conventional risk models (such as the Framingham Risk Score ^29^, Pooled Cohort Equations ^27,28^, QRISK ^27^).
- Reclassification of risk categories often leading to recommendations related to risk-reducing treatments like statins. ^29–31^

While the data strongly suggest CHD PRS, by refining risk estimates, may improve patient outcomes, clinical utility through randomized clinical trials has yet to be conclusively established. We anticipate this is the future direction of PRS studies, and a number of clinical trials are underway. ^32^

### Box 3

**Current breast cancer PRS and their potential uses**

Many of the most recent and most predictive PRS for breast cancer include a smaller number of variants (usually in the 100-1,000s), possibly owing to a less polygenic architecture and more low-frequency variants having greater effect; however, scores composed of millions of variants also exist. Most of the latest PRS have been developed using GWAS summary statistics and data from the Breast Cancer Association Consortium (BCAC), using variants passing genome-wide significance (lead SNPs) or methods including stepwise or penalized regressions on individual-level genotypes ^33^, Pruning + Thresholding ^25^, and Ldpred ^26^. In contrast to CHD, genetics is commonly used to measure and understand breast cancer risk vis-a-vis *BRCA1*/*2* mutation testing; however, routine screening for breast cancer is often performed in older women using mammography and non-genetic risk prediction tools already exist. As such there has been active research into the value of capturing the polygenic risk of common variation on breast cancer, with multiple potential clinical uses and considerations:

- Multiple PRS exist to predict risk for specific subtypes of breast cancer (e.g. ER positive/negative, luminal, and triple negative^33,34^), which could be used to stratify patients for more beneficial treatments, and may have prognostic implications.
- PRS can be combined with existing non-genetic risk prediction models (combining risk factors such as age, family history, mammographic density, hormone replacement therapy) and improve the prediction of incident breast cancer risk. ^35–42^
- Breast cancer has been linked to several rare high-or moderate-risk penetrant genes (e.g. *BRCA1, BRCA2, PALB2, CHEK2, ATM)* and screening for pathogenic variants in these genes is part of clinical practice.^43^ It has been shown that PRS can provide important stratification of risk among carriers of such pathogenic variants and thus could be further useful for clinical decision making. ^26,40,44–48^

Indeed, the BOADICEA breast cancer risk prediction model includes the effects of common variants (PRS313 ^33^) as well as of other rare pathogenic genetic variants ^40^ and has been implemented in the CanRisk Tool (www.canrisk.org) that has been approved for use by healthcare professionals in the European Economic Area. PRS information has been studied via simulation^49^ and is being tested in risk-based breast cancer screening trials in the US ^50^ and Europe (MyPeBS; https://mypebs.eu).

Poorly designed or described studies call into question the validity of some PRS to predict their target outcome ^51,52^, and relatively few studies have benchmarked multiple scores performance externally. At present, there are no uniformly agreed best practices for developing PRS, nor widely adopted standards or regulations sufficiently tailored to assess the eventual clinical readiness of a PRS. There are rapidly emerging applications of PRS that further compound the heterogeneity in reporting, e.g. using PRS as tools for testing gene *x* environment interactions or shared etiology between diseases. ^53–56^ This in itself is not an issue, but the rapid evolution in both the methodological development and applications of PRS make it challenging to compare or reproduce claims about the predictive performance of a PRS for a specific outcome when studies are not properly documented. These deficiencies are barriers to PRS being interpreted, compared, and reproduced, and must be addressed to enable the application of PRS to improve clinical practice and public health.

Frameworks have been developed to establish standards around the transparent, standardized, accurate, complete, and meaningful reporting of scientific studies. In 2011, an international working group published the Genetic Risk Prediction Studies (GRIPS) Statement, a reporting guideline for the study of risk prediction models that include genetic variants, from genetic mutations to gene scores.^57^ This guideline is analogous to guidelines developed for observational epidemiological studies (STROBE^58^) and genome-wide association studies (STREGA^59^), and is in line with the reporting guideline for multivariate prediction models (TRIPOD ^60^). Adherence to reporting statements has been low, and the same holds for GRIPS. One of the reasons might be that researchers feel that GRIPS inadequately addresses PRS, where there has been rapid evolution in score development methodology in recent years. Researchers are frequently uncertain as to what precisely should be reported for a PRS study to be assessed as rigorous, reproducible, and ultimately translatable, especially with the increased push for data availability and transparency. Most PRS studies follow a prototypical process to PRS development evaluation (**Figure 1**) that can be used as a template for standardizing reporting and benchmarking in the field.

**Figure 1:**
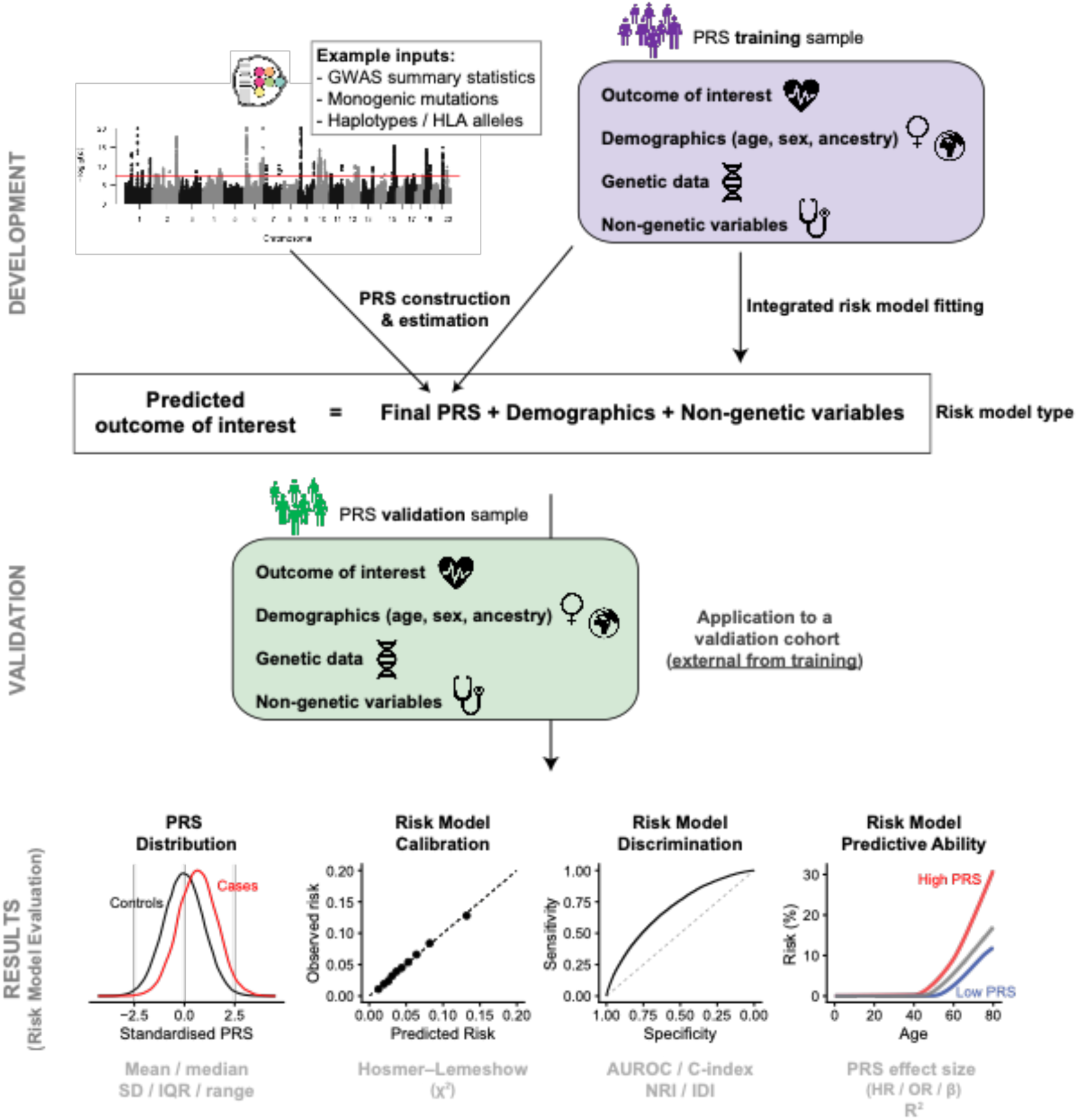
Prototype of PRS development and validation process. Figure 1 displays prototypical steps for PRS construction, risk model development, and validation of performance with select aspects of the PRS-RS guideline labeled in bold text throughout. In PRS development, variants associated with an outcome of interest, typically identified from a GWAS, are combined as a weighted sum of allele counts across variants. Methods for optimizing variant selection for a PRS (PRS construction & estimation) are not shown. The PRS is tested in a risk model predicting the outcome of interest and may be combined with other non-genetic variables (e.g. age, sex, ancestry, clinical variables); collectively these are referred to as risk model variables. After fitting procedures to select the best risk model, this model is validated in an independent sample. The PRS distribution should be described, and the performance of the risk model demonstrated in terms of its discrimination, predictive ability, and calibration. Though not displayed in the figure, these same results should also be reported for the training sample for comparison to the validation sample. In both training and validation cohorts, the outcome of interest criteria, demographics, genotyping, and non-genetic variables should be reported (**Table 1**).

Here, the Clinical Genome Resource (ClinGen) Complex Disease Working Group and the Polygenic Score (PGS) Catalog (**Supplemental Note 1**) jointly present the Polygenic Risk Score Reporting Standards (PRS-RS), a synthesis of an expanded reporting standard for PRS that addresses current research environments with advanced methodological developments to inform clinically meaningful reporting on PRS development and validation in the literature with an emphasis on reproducibility and transparency throughout the development process. Additional methods are detailed in **Supplemental Note 2**.

**Table 1.** Polygenic Risk Score Reporting Statement (PRS-RS)

| Reporting Standard |  | Description |
| --- | --- | --- |
| **Background** | **Study Type** | Specify whether the study aims to develop and/or validate a PRS. When externally validating or combining previously published PRS or integrated risk model, include identifier(s) of original PRS (PMID, PGS Catalog ID). |
|  | **Risk Model Purpose & Predicted End Outcome** | Specify what the risk model is intended to predict and the purpose:<br>Will it be used in risk prediction, diagnostic, prognostic, or therapeutic modalities?<br>What is the end outcome predicted by the risk model? If the predicted outcome is a clinical feature or endpoint within a specific disease, state the disease.<br>What current models for risk prediction are available, if pertinent? |
| **Study Population and Data**<br>*Many risk score studies involve multiple populations and cohorts that can be used in different stages of PRS and risk score development and evaluation. Each of the populations used (e.g. training, validation, subgroup analysis samples) in the manuscript should be defined using this common set of descriptors.* | **Study Design & Recruitment** | For each of the data sets describe the study design (e.g., cohort, case-control, cross-sectional), eligibility criteria, recruitment period and setting (e.g. method and years), and follow-up. State whether the data are primary or secondary data. If secondary analysis, include the full reference to the original study. |
|  | **Participant Demographic and Clinical Characteristics** | Include the distribution of demographic information in each data set (and the combined total if relevant) used to generate a single risk model (whether a single sample set, or the summary of combined samples) including the mean, standard deviation and range. This should at minimum include age, sex and any other characteristics relevant to describe the study population or the performance of the model. Provide demographics stratified by case/control status, if applicable. |
|  | **Ancestry** | Include the ancestral background distribution of each sample population used during PRS development and validation, and the data source of this ancestry information (e.g., self-report, genotyping). Ancestry information should be reported using the standardized framework developed by the NHGRI-EBI GWAS Catalog and ideally include detailed information beyond this when available. When combining samples from multiple studies, aggregate ancestral distribution information is sufficient. The method of ancestry inference should be provided. Genomic methods are preferred, such as principal component analysis. |
|  | **Genetic Data** | Provide method for acquiring genetic information (e.g. sequencing, genotyping) in each sample, including information about genome build and technical details of the assay. If imputing, specify the imputation panel and give ancestry information if not a standard imputation panel. Report any relevant quality control, including imputation quality filters to exclude low quality imputed SNPs. If parameters were selected from another study, include reference (PMID, GWAS Catalog ID). |
|  | **Non-Genetic Variables** | Define any non-genetic variables that were included in the risk model, provide variable definitions and measurement (e.g., assay, ICD codes, e-phenotyping algorithms, chart review, self-report). Indicate the scale of each variable, e.g. dichotomous, continuous, categorical, or ordinal. Explicitly state which variables are included in the final model. |
|  | **Outcome of Interest** | Define the predicted outcome of interest. If the predicted outcome is a clinical feature or endpoint within a specific disease, provide the criteria used to define that disease membership. Include details on how information was ascertained (e.g., ICD codes, e-phenotyping algorithms, chart review, self-report). Transformation of continuous data into binary, ordinal, or categorical outcomes should be detailed with justification. Authors should explicitly state the number of cases and controls. |
|  | **Missing Data** | State explicitly how missing data were handled for all variables included in the model. If imputation was utilized, include detailed of the approach used and any subsequent filtering or post-processing. |
| **Risk Model Development & Application**<br>*Describe the relevant methods used to form the final PRS and/or risk model. Samples used in* | **Polygenic Risk Score Construction & Estimation** | Describe how genetic data were included in the PRS. Authors should detail criteria used to determine inclusion in the model for all variants. Define how the variants were selected, weighted and combined into a single score. If the PRS was derived from another study include the reference (PMID, PGS Catalog Score ID). |
|  | **Risk Model Type** | Detail statistical methods used to estimate risk, either relative or absolute, from the continuous risk score distribution. Detail if risk is cumulative or cross-sectional, as well as the appropriate comparison groups if relative risk presented. Report time until predicted risk (e.g. 5-year, 10-year, lifetime). In a relative hazard model, the study period or follow up |
| *this stage of the analysis should be referred to as "Score Development" or "Training" samples, and be described according to the items in the <u>Study Populations</u> section.* |  | time may be used. In an absolute risk model, state the time until predicted event and the prevalence/incidence of the predicted outcome in the general population. |
|  | **Integrated Risk Model(s) Description and Fitting** | State the procedure utilized to develop the risk models that includes non-genetic and/or genetic variables other than the PRS. If the model(s) was selected for optimal performance, describe measures used to assess performance. Explicitly state all variables used in each risk model. |
| **Risk Model Evaluation**<br>*Outline the results and procedures utilized to evaluate the risk model, specifying internal or external validation. Performance results should be described for both the development and validation samples. Specify if the application of the risk model differs between the development and validation samples.* | **PRS Distribution** | Include a general description of the distribution of the PRS. This details the continuous distribution output directly from the risk score calculation. |
| | **Risk Model Predictive Ability** | Describe and report any metrics of overall performance (proportion of variance explained; $R^2$) and estimates of risk (such as odds or hazards ratios from regression models) used to evaluate the PRS and/or risk models. Describe the set of genetic/non-genetic variables included in the analysis. |
|  | **Risk Model Discrimination** | Describe and report metrics used to assess the discrimination of evaluated risk models and whether any non-genetic variables were included beyond a PRS in this analysis. Common metrics include the area under the ROC or Precision-Recall (AUROC/AUPRC) and the Concordance statistic (C-index) for survival models. Evaluation of potential clinical utility of models requires also evaluating tail-based measures, such as proportions of populations and cases exceeding specified clinically relevant risk thresholds and measures of reclassifications (e.g. NRI) at such thresholds for comparison of models. |
|  | **Risk Model Calibration** | Describe and report metrics used to assess the calibration of evaluated risk scores and models. Describe the set of genetic/non-genetic variables included in the analysis. |
|  | **Subgroup Analyses** | Subgroup size, demographics and clinical characteristics should be given. Relevant evaluation methods and measures (distribution, predictive ability, discrimination, calibration) should be described for each subgroup analysis. |
| **Limitations and Clinical Implications**<br>*Discuss the broader context of the study and risk model.* | **Risk Model Interpretation** | Summarize the risk models in terms of what they predict, how well, and in whom. Explicitly mention the performance of the PRS and/or combined risk model in comparison to conventional risk models. Conventional risk models might include demographic (age, sex), disease-specific risk factors, and/or family history of disease. |
|  | **Limitations** | Outline limitations of the study with relevance to the results, discuss the impact of these limitations on the interpretation of the risk model and any downstream replication efforts needed. Common considerations include: study design restrictions, use of a surrogate outcome, ascertainment biases, the distribution of participant-level traits (ancestry, age, comorbidities), accuracy/specificity of outcome data, and any statistical considerations. Make note of and discuss the impact of any unknown reporting items from previous sections. |
|  | **Generalizability** | Discuss the intended target groups or populations this score may be applied to and explicitly address any issues with generalizability beyond the included populations. Discuss whether the study externally validates the score and/or model, or if the sample is limited with respect to ancestry, age, or other variables. |
|  | **Risk Model Intended Uses** | Discuss whether there is an intended clinical use or utility to the risk model. If so, discuss the "clinic readiness" and next steps with respect to the interpretation, limitations, and generalizability of the model. Discuss how the predictive ability of the model is compared against current standard of care or other published work (such as existing PRS) on predicting the outcome of interest. |
| **Data Transparency and Availability** |  | Information sufficient to calculate the PRS and the risk model(s) on external samples should be made freely available. For genetic variables this would include information about the variants (e.g., rsID, chromosomal location, effect allele, and the effect weight) that comprise the score; PRS with this information should be deposited in the PGS Catalog for findability and to promote re-use and comparison with other established scores. Weights for non-genetic variables should also be provided to make the risk model calculable. |

## The ClinGen-PGS Catalog Polygenic Risk Score Reporting Standards (PRS-RS)

The PRS-RS is a set of standard items specifying the minimal criteria that need to be described in a manuscript in order to accurately interpret a PRS and reproduce results throughout the PRS development process, briefly illustrated in **Figure 1**. ^61^ It applies to both PRS development and validation studies that aim to predict disease onset and prognosis, as well as response to therapies; however, other research uses of PRS have overlapping steps that should be reported similarly. **Table 1** presents the full PRS-RS, in which criteria are organized into key components along the developmental pipeline of PRS for clear interpretation and to encourage their documentation from the inception of the study well before publication.

### Reporting on risk score background

The development and validation of a PRS tests a specific hypothesis with a defined outcome and study population. Therefore, authors should define *a priori* the *study type* (e.g. development and/or validation), *purpose* (e.g. risk prediction vs. prognosis) and *predicted end outcome* (e.g. CHD) in enough detail to understand why the study population and risk model selected are relevant (e.g. the value for CHD risk stratification and primary prevention is highest in younger individuals compared to those over 80 who have accumulated risk over a lifetime). As the PRS-RS is focused on clinical validity and implementation, authors must outline the study and appropriate outcomes to understand what risk is measured, what the purpose of measuring risk would be, and why this purpose may be of clinical relevance. In order to establish the internal validity of a study, authors should use the appropriate data needed to address the intended purpose (e.g. prediction of incident disease vs. prognosis), with adequate documentation of dataset characteristics to understand nuances in measured risk.

### Reporting on study populations

The applicability of any risk prediction to an external target population (the “who, where, and when”) depends on its similarity to the original study populations used to derive the risk model. Therefore, authors need to define and characterize the details of their study population (e.g. *study design and recruitment*), and describe *participant demographics* for key variables (such as age and sex) and *ancestry*. Importantly, there are often inconsistent definitions and levels of detail associated with ancestry, and the transferability of genetic findings between different racial/ethnic groups can be limited. ^1,9,62^ It is therefore essential for authors to provide a detailed description of participants’ genetic ancestry - including how ancestry was determined – using a common controlled vocabulary where possible (e.g. tables 1 and 2 of the standardized framework developed by the NHGRI-EBI GWAS Catalog^1^). While age, sex and ancestry are the most universally relevant characteristics, authors should provide a sufficient level of detailed criteria for defining all the relevant factors used in the risk model (*non-genetic variable(s)*) and the *outcome of interest*. This is particularly important if they are included in the final risk model and should accompany information about how the population was genotyped (*genetic data*).

### Reporting on risk model development

There are currently several methods that are commonly used to select variants and fine-tune weights that constitute the PRS. ^7,16–18,61^ For methods using individual-level genomic data, the original source study should be cited or the assays and quality control described in detail. Methods using GWAS summary statistics should clearly cite the relevant GWAS(s) (preferably using unique and persistent study identifiers from the GWAS Catalog (GCSTs; ^63^). As the performance and limitations of the combined risk model are dependent on methodological considerations, authors must provide complete details including the method used and how variants are selected and combined into a single PRS (*PRS construction & estimation*). Apart from the genetic data in the model, authors should also describe the defining criteria for other demographic and non-genetic predictors (*non-genetic variables*) included in the model. Often authors will iterate through numerous models to find the optimal fit. In addition to the estimation methods, it is important to detail the *integrated risk model fitting* procedure, including the measures used for final model selection. Translating the continuous PRS distribution to a risk estimate, whether absolute or relative, is highly dependent on assumptions and limitations inherent to the specific data set utilized. When describing the *risk model type*, authors should detail the time scale employed for prediction or the study period/follow-up time for a relative hazard model. Additionally, if relative risk is estimated, the reference group should be well described. These details should be described for the training set, as well as validation and sub-group analyses.

### Reporting on model parameters

Authors should report estimates for all evaluated models, not only the methods behind decision-making, equipping readers with the information necessary to gauge the relative value of an increase in performance against other trade-offs (*data transparency and availability*). The underlying PRS (variant alleles and derived weights) should be made publicly available, preferably through direct submission to an indexed repository such as the PGS Catalog, to enable others to reuse existing models (with known validity) and facilitate direct benchmarking between different PRS for the same trait. The current mathematical form of most PRS—a linear combination of allele counts—facilitates clear model description and reproducibility. Future genomic risk models may have more complex forms, e.g. allowing for explicit non-linear epistatic and gene-environment interactions, or deep neural networks of lesser clarity. It will be important to describe these models in sufficient detail to allow their implementation and evaluation by other researchers and clinical groups.

### Reporting on risk model evaluation

We recommend that authors provide summary information of the *risk score distribution* to aid in model interpretation. The *risk model’s predictive ability, calibration*, and *discrimination* should also be assessed and described with common descriptions including the risk score effect size, variance explained (R^2^), reclassification indices, and metrics like sensitivity, specificity, positive predictive value (PPV), and negative predictive value (NPV). The *risk model calibration* and *discrimination* should be described for all analyses, although their estimation and interpretation are most relevant for the PRS validation sample. It is imperative for the PRS and expanded risk models to be evaluated on a population that is external (e.g. independent, non-overlapping) to the individuals in the study population. The ability of the risk model to classify individuals of interest (*risk model discrimination*) can commonly described and presented in terms of the AUROC, AUPRC or C-index. Any differences in variable definitions or performance discrepancies between the training and validation sets should be described.

### Reporting on interpretation

By explicitly describing the *risk model’s interpretation* and outlining potential *limitations* to the *generalizability* of their model, authors will empower readers and the wider community to better understand the risk score and its relative merits. Authors should justify the clinical relevance and *risk model intended uses*, such as how the performance of their PRS compares to other commonly used risk models, or previously published PRS. This may also include comparisons to other genetic predictors of disease (e.g. mutations in high/moderate risk genes associated with Mendelian forms of the disease), family history, simple demographic models, or conventional risk calculators (see Box 2 and 3 for disease-specific examples). What indicates a “good” prediction can differ between outcomes and *intended uses*, but should be reported with similar metrics to what is described in the evaluation section.

**Supplemental Note 3** provides reporting considerations in addition to the minimal reporting framework in **Table 1**. Authors intending downstream clinical implementation should aim for the level of transparent and comprehensive reporting covered in both **Table 1** and **Supplemental Notes**, especially those related to discussing the interpretation, limitations, and generalizability of results. The proper reporting of PRS development and performance can also have implications for seeking regulatory approval of the PRS as a clinical test. Though not a comprehensive list of regulatory requirements, we highlight aspects of PRS-RS that would be considered evidence of analytical and clinical validity from the College of American Pathologists (CAP) and the Clinical Laboratory Improvement Amendments (CLIA) perspective (**Supplemental Table 1**). CAP and CLIA approvals are additional incentives for reporting adherence of researchers wishing to translate their work, as well as a caution for researchers wishing to avoid unintended uses of their findings. Lastly, we reiterate the need for both methodological and data transparency and encourage deposition of PRS (variant-level information necessary to recalculate the genetic portion of the score) in the PGS Catalog (www.PGSCatalog.org; ^64^), which provides an invaluable resource for widespread adoption and distribution of a published PRS. The PGS Catalog provides access to PGS and related metadata to support the FAIR principles of data stewardship ^19^, enabling subsequent applications and assessments of PGS performance and best practices (see **Supplemental Table 2** for a description of the metadata captured in the Catalog and it’s overlap with the PRS-RS).

## Using the PRS-RS and PGS Catalog to improve PRS research and translation: transparency and open data

We surveyed 30 publications (selecting for a diversity of disease domains, risk score categories, and populations) to understand how the information in the PRS-RS is presented and displayed as part of the larger iterative process to clarify and improve field definitions. For 10 of these publications, we provide detailed annotations using the final field definitions (**Supplemental Table 3**) and use these annotations to illustrate the detail necessary for each PRS-RS item (further described in **Supplemental Note 3**). The heterogeneity in the PRS reporting we observed in this pilot highlights a series of challenges. Critical aspects of PRS studies, including ancestry, predictive ability, and transparency/availability of information needed to reproduce PRS, were frequently absent or reported in insufficient detail. This underscores the need for PRS-RS to clearly and specifically define meaningful aspects of PRS development, testing, and intended clinical use. However, these deficits in reporting are not unique to PRS; previous reports of underreporting have found that 77% of GWAS publications in 2017 did not share summary statistics ^65^ and 4% of GWAS do not report any relevant ancestry information ^1^. In line with the push towards a culture of reproducibility and open data in genomics, we as the ClinGen Complex Disease Working Group and PGS Catalog joined to create a set of reporting standards (**Table 1**) specifically tailored to PRS research based on multidisciplinary and international expert opinion for tailoring previous standards.

Researchers using PRS-RS may identify fringe cases that are inadequately captured by these reporting items, as we have modeled our guideline on prototypical steps for PRS development (**Figure 1**) for flexibility. While we anticipate the field may further change as novel methods and technologies are generated, the PRS-RS items can be further expanded and adapted to encompass novel considerations. By updating previous standards, drawing upon current leaders in the field, and tailoring the framework to common barriers observed in recent literature, we aim to provide a comprehensive and pragmatic perspective on the topic. In line with previous standards, PRS-RS includes elements related to understanding the clinical validity of PRS and consequent risk models. Items such as *predicted end outcome* and *intended use* bookend our guideline with the intended clinical framing of PRS reporting. In addition, we have modeled the guideline by steps in experimental design, from hypothesis to interpretation, to more clearly emphasize the significance of the intended use case in defining what needs reported and inform documentation throughout the process. As a reference, we have also included a guide to where PRS-RS items should be reported in a manuscript in **Supplemental Table 4**). These expansions will further facilitate the curation and expert annotation of published PRS as we move towards widespread clinical use.

While the scope of our work encompasses clinical validity, it does not address the additional requirements needed to establish the clinical or public health utility of a PRS, such as randomized trials with clinically meaningful outcomes, health economic evaluations, or feasibility studies. ^66^ In addition, the translation of structured data elements into useful clinical parameters may not be direct. A relevant example is that the disease case definitions utilized in training or validation in any particular PRS study may deviate (sometimes substantially) from those utilized in any specific health system, for example CHD symptoms commonly include angina (chest pain), whereas PRS are frequently trained on stricter definitions excluding angina. In addition, the definitions used for race/ancestry as outlined in the PGS and GWAS Catalog^1^ may also differ from structured terms used to document ancestry information in clinical care, in which case consistent mappings, and potentially parallel analyses, may be necessary to translate from genetically determined ancestries to those routinely used in clinical care. Such translation issues potentially limit generalizability to target populations and warrant further discussion, and we reiterate the need for authors to be mindful of their intended purpose and target audience when discussing their findings. Nevertheless, we have aided authors’ understanding of potential translational barriers by considering current CAP/CLIA analytical and clinical validity evidence requirements of peer-reviewed literature to ensure PRS-RS has value in informing later steps of the clinical translation spectrum, including clinical utility (**Supplemental Table 1**). Finally, while the principles of this work are clear, its scope does not include the complex commercial restrictions, such as intellectual property, that may be placed on published studies regarding the reporting or distribution of PGS, or the underlying data thereof. We hope this work will inform downstream regulation and transparency standards for PRS as a commercial clinical tool.

The coordinated efforts of the ClinGen Complex Disease Working Group and PGS Catalog provide a set of compatible resources for researchers to deposit PGS/PRS information. The PGS Catalog (www.PGSCatalog.org) provides an informatics platform with data integration and harmonization to other PGS as well as the source GWAS study through its sister platform, the GWAS Catalog. ^64^ In addition, it provides a structured database of scores (variants and effect weights) that can be reused, along with metadata requested in the PRS-RS. With these tools, PRS-RS can be mandated by leading peer-reviewed journals and, consequently, the quality and rigor of PRS research will be elevated to a level which facilitates clinical implementation. We encourage readers to visit the ClinGen complex disease website (https://clinicalgenome.org/working-groups/complex-disease/) for any future changes or amendments to the reporting guideline.

While we have provided explicit recommendations on how to acknowledge study design limitations and their impact on the interpretation and generalizability of a PRS, future research should attempt to establish best practices to guide the field. Moving forward, supplemental frameworks should be developed for the reporting of new methods, such as deep learning, as well as requirements for clinical utility and readiness. Taken together, PRS-RS facilitates the rapid emergence of polygenic risk scores as potentially powerful tools for the translation of genomic discoveries into clinical and public health benefits, and provides a framework for PRS to transform multiple areas of human genetic research.

## Data Availability

N/A

## Acknowledgements

We would like to acknowledge the input of the ClinGen Complex Disease Working Group members, including Carlos D. Bustamante, Michelle Meyer, Frank Harrell, David Kent, Peter Visscher, Tim Assimes, Sharon Plon, and Jonathan Berg. We would also like to thank Diane Durham for her editorial support and Angela Paolucci for her administrative support in the preparation and submission of this manuscript.

ClinGen is primarily funded by the National Human Genome Research Institute (NHGRI), through the following three grants: U41HG006834, U41HG009649, U41HG009650. ClinGen also receives support for content curation from the Eunice Kennedy Shriver National Institute of Child Health and Human Development (NICHD), through the following three grants: U24HD093483, U24HD093486, U24HD093487. The content is solely the responsibility of the authors and does not necessarily represent the official views of the National Institutes of Health. Additionally, the views expressed in this article are those of the author(s) and not necessarily those of the NHS, the NIHR, or the Department of Health. Research reported in this publication was supported by the National Human Genome Research Institute of the National Institutes of Health under Award Number U41HG007823 (EBI-NHGRI GWAS Catalog, PGS Catalog). In addition, we acknowledge funding from the European Molecular Biology Laboratory. Individuals were funded from the following sources: MIM was a Wellcome Investigator and an NIHR Senior Investigator with funding from NIDDK (U01-DK105535); Wellcome (090532, 098381, 106130, 203141, 212259). MI, SAL, and JD were supported by core funding from: the UK Medical Research Council (MR/L003120/1), the British Heart Foundation (RG/13/13/30194; RG/18/13/33946) and the National Institute for Health Research (Cambridge Biomedical Research Centre at the Cambridge University Hospitals NHS Foundation Trust). SAL is supported by a Canadian Institutes of Health Research postdoctoral fellowship (MFE-171279). JD holds a British Heart Foundation Personal Chair and a National Institute for Health Research Senior Investigator Award. This work was also supported by Health Data Research UK, which is funded by the UK Medical Research Council, Engineering and Physical Sciences Research Council, Economic and Social Research Council, Department of Health and Social Care (England), Chief Scientist Office of the Scottish Government Health and Social Care Directorates, Health and Social Care Research and Development Division (Welsh Government), Public Health Agency (Northern Ireland), British Heart Foundation and Wellcome.

## Author Contributions

HW, CT, MAI, IJK, CS, RR, KV, KK, KABG, PK, and GLW conducted the literature review of reporting practices. HW, SAL, CT, MIA, JWO, IJK, RR, DB, EV, MIM, ACA, DFE, RAH, AVK, NC, CK, KE, EMR, MCR, KEO, MJK, ACJWJ, KABG, PK, JALM, MI, and GLW provided feedback on the reporting framework. HW, SAL, CT, MAI, JWO, IJK, AVK, JND, HP, EMR, MR, ACJWJ, KABG, PK, JALM, MI, and GLW wrote and edited the manuscript text and items.

