## Supplemental Notes for "Improving reporting standards for polygenic scores in risk prediction studies"

### 1. Description of the author groups and project goals

#### 1.1 ClinGen Complex Disease Working Group

The working group, founded by ClinGen in November 2018, composed of more than thirty experts with epidemiological, statistical, disease-domain specific, implementation science, actionability, and ELSI interests in polygenic risk score application. Members met twice a month to discuss current research, best practices, and limitations within their respective areas of expertise. As a result of these meetings, the workgroup decided to update previous genetic risk-score reporting standards [1] to current PRS practices. This aim was finalized at the NHGRI Genomic Medicine XII: Genomics and Risk Prediction meeting in May 2019 with input from the external scientific community in terms of mission, scope, and long-term objectives of the working group. Current descriptions of workgroup members and goals are available at: https://clinicalgenome.org/working-groups/complex-disease/

#### 1.2 The Polygenic Score (PGS) Catalog

The PGS Catalog was founded in 2019 by researchers at the University of Cambridge UK, European Bioinformatics Institute (EMBL-EBI) and Baker Institute, and developed as a sister resource to the NHGRI-EBI GWAS Catalog [2,3]. Its goal is to provide an open database of PGS and relevant metadata, so that published PRS/PGS can be distributed, applied, and evaluated in a rigorous and replicable manner in both research and clinical settings. It reports key information about how a PGS has been developed (e.g., variant selection and computational methods), information about the specific datasets used for PGS development and evaluation (e.g., sample size, ancestry, phenotype description), as well as the performance metrics reported during PGS evaluation (e.g., effect sizes, covariates, and/or classification metrics). These data are represented in a schema that links the Scores, Samples, and Performance Metrics presented in each PGS publication. The PGS Catalog is available at www.PGSCatalog.org; additional descriptions of the project, development, methods, full descriptions of the representation schema, along with links for PGS submission can be found in the documentation (www.pgscatalog.org/about/) and are described in a separate publication about the Catalog. [4]

### 2. PRS-RS Development Process

#### 2.1 Framework construction

The Polygenic Risk Score Reporting Statement (PRS-RS) was developed through the in iterative phases utilizing previous standards, expert opinion adaptation of previous genetic risk study reporting standards to fit the prototypical steps in PRS development and evaluation. First, the entire expert working group created the initial framework draft by adapting previous genetic risk-score reporting standards (GRIPS, 2011[1]) to current PRS methodologies. We primarily relied on expert opinion for this initial expansion and incorporated principles guided by the PICOT framework [5], which is used to compare heterogeneous clinical trial outcomes, since heterogeneity was a major anticipated concern in PRS reporting. Our revisions focused on eliciting the individual components from previous standards that experts deemed independently important for transparent interpretation and reproducibility of a risk score, especially with regard to any downstream clinical application. We expanded the original GRIPS checklist of 25 items to 44 unique items, of which 33 items were needed for both training and validation cohorts. The majority of these additions were added to explicitly list discrete elements within an individual GRIPS checklist item if those elements were determined by the work group to have significant impact in the interpretation of a PRS in terms of either analytic validity, clinical validity, or clinical utility. The PICOT framework did not add items to the reporting guidelines, but we did confirm that PICOT concepts were represented in the reporting guideline to facilitate downstream applications of comparing heterogeneous outcomes.

#### 2.2 Testing PRS-RS on existing publications

We used the PRS-RS checklist to beta test published original research articles on polygenic risk-score development or validation as a measure of pragmatism and clarity. Thirty-five papers were initially collected via the snowball sampling search based on their use of the term "polygenic risk score" and their research in human populations in preparation for the NHGRI Genomic Medicine XII meeting. Five papers were excluded from the review because they were not original articles, did not develop or validate a PRS, duplicated a previous study, or did not use genetic loci to construct their risk scores. Included articles spanned a variety of disease domains including Alzheimer's disease, asthma, breast cancer, cerebrovascular event, colon cancer, coronary heart disease, depression, fracture risk, Parkinson's disease, prostate cancer, and schizophrenia. In addition, articles were selected for variety in the risk score category (development vs. external validation; diagnostic vs. prognostic). Article references are available in the supplement. The majority of papers (25/30) were predicting risk of developing disease with a few characterizing prognostic outcomes. Nearly half of the papers (13/30) developed a novel risk score, while the other half either externally validated a previously published risk score (9/30) or both developed and externally validated the risk score (6/30). Two manuscripts modified a previously published score. The composition of the final published risk scores were limited to genetic variables for the majority of papers (25/30), with only five producing an integrated risk score.

Two independent reviewers assessed each article using the draft PRS reporting framework. A 10-person volunteer subgroup of the larger working group met bi-weekly to resolve inter-reviewer discrepancies. If the subgroup was unable to reach a consensus, one of four expert reviewers from the working group was assigned to resolve discrepancies in a third review. This pilot of the reporting guideline on published PRS revealed pragmatic areas for revision. Similar items were combined if they did not individually contribute meaningful concepts for PRS interpretation. Items were removed if they did not contribute to overall interpretation of the risk-score performance or target application. Definitions were expanded and revised to address inconsistencies in inter-reviewer interpretation due to heterogeneous and vague reporting in the literature. Items were kept as discrete items if we observed substantially missing or insufficiently detailed reports on these items in the literature for transparency. When applicable, updated methodology was also included in definitions. Finally, supplemental considerations were created to address fringe cases (*see Explanation and Considerations below*). Proposed reporting guideline revisions were ratified in monthly calls with the entire workgroup. This resulted in an initial 33 item reporting guideline that organized the reporting items into manuscript sections.

Papers were curated once again using the 33-item draft reporting guideline. This process revealed that the formatting of a reporting guideline by manuscript section created redundancy and confusion. It was decided that reformatting the guideline to reflect experimental steps of PRS development and evaluation both streamlined the guideline, reducing it to a final 23 items (**Table 1**), and more clearly illustrated the need for hypothesis-driven reporting. This rearrangement was largely guided by harmonization with the PGS catalog, which models these experimental steps.

The majority of papers (25/30) were predicting risk of developing disease with a few characterizing prognostic outcomes. Nearly half of the papers (13/30) developed a novel risk score, while the other half either externally validated a previously published risk score (9/30) or both developed and externally validated the risk score (6/30). Two manuscripts modified a previously published score. The composition of the final published risk scores were limited to genetic variables for the majority of papers (25/30), with only five producing an integrated risk score. As an example, 10 of these papers were curated with the final 23-item reporting framework in **Supplemental Table 3**.

#### 2.3 PRS-RS harmonization with PGS Catalog

For this resource harmonization step, two curators mapped reporting fields from the PGS Catalog onto the final PRS-RS guidelines. When possible, similar terminology was adopted between the two resources. A subset of fields in the PGS Catalog differ from PRS-RS due to restrictions in preserving integrity of the data infrastructure. The analogous ClinGen reporting item to each PGS Catalog field is presented in Supplementary Table 2.

The goals of the PGS Catalog align with ClinGen with slight differences in how the data is represented in the Catalog (link). Overall, there is a good agreement between the PRS-RS and PGS Catalog representation schema (field by field mapping outlined in **Supplemental Table 2**), particularly with respect to how study participants are described. The five reporting items in the PRS-RS that are not present in the PGS Catalog include descriptions, goals, limitations and intended uses of PRS predictions and implementation that are not essential to the Catalog's goal of indexing available published PGS with the metadata essential for interpretation and reproducibility. PRS described using the PRS-RS items contain sufficient detail for their addition to the PGS Catalog, as such we recommend that authors describe scores using the PRS-RS and submit them to the PGS Catalog upon publication.

### 3. Explanation and Considerations

Here we include a detailed explanation of PRS-RS items, including considerations specific to each item regarding limitations and best practices.

#### 3.1 Risk Model Background

This section details the components necessary in framing the development and potential downstream utility of a polygenic risk score. Authors should have a clear vision of how this score would be used, including the appropriate target population and clinical end outcome. By contextualizing the motivation behind the PRS development and/or validation, readers will be better able to judge the appropriateness of relevant statistical methods and study population. We recognize that the development and use of a polygenic score can be motivated for a range of purposes unrelated to clinical utility. While PRS-RS is written with an eye towards clinical utility, nearly all of the fields are applicable to other purposes as well, including gene by environment interactions and shared etiology between traits as it is equally important to detail the definitions and methods utilized for proper interpretation. The main difference will be the **Risk Model Background** (**Section 3.1**), specifically the *risk model purpose*, and the **Limitations and Clinical Implications** (**Section 3.5**).

**Study Type.** Manuscripts detailing the development of polygenic risk scores can vary in the stage along the pipeline, from effect size estimation to external validation. Authors must be clear in the scope of their manuscript, specifically whether effect sizes were estimated directly from individual-level data as part of this manuscript, built upon a previously-published genome-wide association study (GWAS), or if the study seeks to externally validate a previously-published PRS. If building upon previous GWAS or PRS, authors should include the appropriate identifier(s) of data utilized (PMID, GWAS Catalog ID, PGS Catalog ID).

**Risk Model Purpose & Predicted End Outcome.** It is important for authors to determine *a priori* the ultimate target of the risk score, especially in the context of downstream clinical utility. This includes specifying what the risk score is intended to predict and the purpose of this risk score, such as risk prediction, diagnostic, prognostic, or therapeutic modalities (or a combination). If the model is trained on a different measure than the intended end outcome, authors should justify this discrepancy and provide evidence for the appropriateness of this decision. For example, a PRS is developed with the clinical end outcome of type II diabetes diagnosis. However, investigators do not have access to T2D diagnoses in their training dataset. Instead, they use HbA1c measures to train their PRS, which is then externally validated on a dataset with T2D information. Authors should detail the caveats in using HbA1c, a continuous measure of glycemic control, to diagnose individuals based on genetics, including the reliability of this measure with respect to demographics (such as race/ethnicity, age, sex, etc.). This may include possible population stratification, as individuals without diabetes mellitus have been shown to have different levels of HbA1c by self-identified race/ethnicity. [6] Authors should also detail what indicates a “good” prediction for their specified outcome and purpose, preferably alongside other clinical risk models and standard practice. By contextualizing the PRS within current practice and the state of the field, authors better facilitate the downstream evaluation and adoption of their clinical tool.

#### 3.2 Study Populations

As with any epidemiological study, it is important to fully understand the study population and its relevance to a target population for clinical utility. By fully describing the sample characteristics and how data was handled, authors provide valuable insight for the downstream utility of the PRS and how/why performance may differ between populations. Many genetic studies involve multiple populations and cohorts along the development and validation pipeline and it is important for authors to describe in as much detail as possible each of these samples. This includes different cohorts, as well as different stages such as training, validation, and subgroup analyses within the manuscript. Variable definitions should be consistent across different cohorts and stages with documented harmonization protocols.

**Study Design and Recruitment.** The performance, interpretation, and downstream clinical utility of a polygenic risk score is highly dependent on the characteristics of its study population. Therefore, it is imperative for authors to describe the study design and recruitment processes with as much detail as possible. The study design determines the relevance of the statistical model, and therefore must be described, such as the type of sample (e.g., cohort, case control, cross sectional) and recruitment details (e.g. method and years). These characteristics will inform whether the predicted clinical end outcome is defined by incidence or prevalence. If applicable, justify the use of prevalent disease for your intended use. Additionally, the performance of a PRS should not be determined in a case-control study, as the lack of representative samples limits the calculation of risk. If a case-control study design is utilized, authors should justify their choice and discuss potential limitations. If prevalence is used, authors must describe their reasoning and how that may limit the downstream utility of their PRS in determining future risk.

**Participant Demographics and Clinical Characteristics**. Investigators should describe the distribution of demographic information of their samples, including but not limited to age and sex, to better inform readers as to the relevance of the study population to the target population. In addition to these variables, authors should detail all information relevant to the prediction model's predicted end outcome and intended use, such as whether included patients are early stage, late stage, or pre-symptomatic. As many biomedical outcomes are age-dependent, it is important to know the age distribution of the study population for PRS development and validation. This should include the mean, standard deviation, and range of ages, preferably by case/control status, if applicable. If the intended use of the PRS is for a specific age range, authors should provide additional statistics focused on that age range and representation within the sample population. If longitudinal data is used, authors should specify the age distribution at the beginning of follow-up, as well as any relevant time periods to the predicted outcome of interest with attention to loss-to-follow-up by age. As with age, sex can be an important factor to consider in PRS performance. Therefore, the sex distribution should be described as both the counts and percentages of the total sample. Authors should state if sex was inferred from self-report or genetic information. If applicable, sex distribution should be provided by case/control status. If the study explicitly refers to gender instead of sex, details should be provided to differentiate between the definitions and how they are relevant to study goals. The limitations of self-report should be addressed, such as how self-report more accurately reflects gender identity than biological sex assigned or defined at birth. Additional guidance can be found in the NIH Policy on Sex as a Biological Variable (https://orwh.od.nih.gov/sex-gender/nih-policy-sex-biological-variable).

**Ancestry.** The genetic ancestry of participants has profound consequences for both the performance of a polygenic risk score and its appropriateness for downstream clinical utility in various populations. Because of this, authors should provide extensive ancestry information to contextualize results, preferably by case/control status, if applicable. This should include self-identified race/ethnicity and genetically-determined ancestry. If principal components are used to determine genetic ancestry, plots and interpretation should be provided both with and without available reference panels (such as the 1000 Genomes Project) to allow comparison with external studies. We encourage authors to follow a standardized framework for ancestry as developed by the NHGRI-EBI GWAS Catalog [7] at the minimum. Ethnocultural descriptors should provide information about the underlying genetics or if epidemiologically relevant. For founder populations, the broader genetic background should also be described. Admixed populations should also have the ancestral backgrounds described that contribute to the admixture. If ancestry is not known or not able to be disclosed, authors must explicitly state this and the reasons why in the discussion as a limitation. Geographic location should not be used as a proxy to infer ancestry information and should be explicitly stated as a location, not ancestry.

**Genetic Data.** Authors should detail the method for acquiring genetic information, such as the technology (sequencing versus genotyping), as well as information of genome build and technical details of the assay. If an array was used with imputation, authors need to explicitly describe the imputation process, including population representation on the imputation panel and quality control measures enacted to select SNPs for PRS. All imputation quality filters should be reported to exclude low quality imputation SNPs. Authors should also state if imputed SNPs included in the PRS were experimentally validated. If data acquisition differed across combined samples, these different processes should be described with sample sizes for the subsampled groups. If any of this information or relevant parameters were selected from another study, authors should also include relevant references (e.g. PMID, GWAS Catalog study ID).

**Non-genetic Variable(s).** Many PRS include variables in the prediction models in addition to the genetic variants and their effect sizes. These variables, such as age, sex, race/ethnicity, and measurements of disease specific biomarkers/risk factors (e.g. cholesterol, C-reactive protein, insulin resistance) can have profound influence on the performance of a PRS and therefore must be described in detail, preferably with justification for why the variable might warrant inclusion in the risk model. This includes the inclusion and exclusion criteria for each variable, as well as the data source for that information (e.g. ICD codes, e-phenotyping algorithms, chart review, self-report). Authors should also indicate whether the variable was included as a dichotomous or continuous measure. Authors should also explicitly state which variables are included in the final risk model as not all variables included in the risk model may be retained after model fitting.

**Outcome of Interest.** As previously mentioned in the Risk Score Background section above, it is vital for the authors to define the end outcome targeted with the PRS. If this outcome is a clinical feature or endpoint within a specific disease, inclusion and exclusion criteria need to be defined, as well as how that information was ascertained (similar to clinical variable definitions above). If a dichotomous outcome is utilized, numbers of cases and controls should be stated along with their specific inclusion/exclusion criteria, both overall and by specific subpopulations or important clinical variables. All data transformations (continuous to binary outcomes, normalization) should be detailed to enable reproducibility. For a validation study, authors should describe how the predicted outcome may differ from the original phenotype in the score development, as well as a justification for the use of the outcome despite these differences.

**Missing Data.** Protocols for the handling of missing data should be detailed for all variables used in the development of the PRS, including both genetic and non-genetic factors. If variables were not missing at random, authors should include in the discussion how this may limit the interpretation of their final score as to the representativeness of their model to the target population.

#### 3.3 Risk Score Development and Application

Once readers are well acquainted with the details of how data was created, curated, and cleaned, the next step is understanding the statistical methods and measures utilized in the development of the polygenic risk score.

**Polygenic Risk Score Construction & Estimation.** The construction of a PRS often goes through the following major steps: (1) variant-level effect size estimation, (2) selection of variants to include in PRS, and (3) pooling of variant-level effects, with or without weighting. For each of these steps, authors should describe their methods with detail to enable reproducibility. For step 1, authors should describe the statistical model used to estimate effect sizes, such as a linear or logistic regression, as well as any covariates included in addition to the genetic effects. Training samples and variant selections should be clearly described alongside these computational methods. Other common methods may include snpnet, BLUP-based methods, regularized regression (e.g. LASSO/ridge), or stepwise regression. Whether the effect size refers to allelic or genotypic risk should be stated, as well as mode of inheritance (additive, dominant, recessive, etc.) If a previously published GWAS is used, authors should include the full citation including PMID. For the selection of SNPs (step 2), authors should list any thresholds that were used, such as P-value thresholds, or functional effects, both for the final model and all tested models. Details for pruning using linkage disequilibrium should include the population (reference panel or study population) used to estimate LD, as well as the LD threshold ($r^2$, D'). If effect sizes are reweighted from their original estimates (step 3), procedures should be well-documented and parameters should be justified given the outcome and study population. Such methods may include LDpred, lassosum, meta-scoring approaches, etc.

**Risk Model Type**. After the calculation of the polygenic risk score, the continuous distribution must be translated into estimated risk for study participants. The two ways to assess an individual's risk are absolute versus relative risk, or whether risk is qualified compared to a reference group or whether it is the absolute probability of the event occurring. As many PRS are developed with case-control studies, relative risk is often calculated as the risk of one strata compared to another. Authors should be explicit when describing this relative risk and in defining the reference group. If absolute risk is estimated, authors should be explicit in any assumptions or external data used to calculate this measure. As many PRS are developed to determine the risk of a future event, the time period must be well defined. If a relative hazards model is used, authors should describe the time period and justify their selection with a lens of downstream clinical relevance and utility. If absolute risk is calculated, authors should specify the time to event. Any external information, such as disease prevalence and demographics, used to derive absolute risk should be described and cited. Authors should be careful not to simply report total length of study.

**Integrated Risk Model(s) Description and Fitting.** The selection of the final PRS model can be an iterative process, selecting different subsets of variants and non-genetic variables to optimize prediction accuracy. The metric used to select the optimal model can be dependent on the predicted clinical end outcome (binary, continuous), as well as the potential impact on clinical utility such as maximizing sensitivity or specificity. Authors should detail all procedures used to select the final model, including the statistical metric of accuracy and any other factors such as minimal sets of SNPs. All metrics should be reported in the main text or supplement for all models compared, including calibration and discrimination as applicable. This may include stepwise regression for the integration of non-genetic clinical variables. The formulation of the final risk score should be detailed, including how all variables are coded (genetic and non-genetic). If race, ethnicity or ancestry (REA) is included in the model, authors should specify how this is measured (self-report, genetic) and all statistical methods utilized. When applicable, methods appropriate to admixed ancestries (e.g. African American and Hispanic/Latino populations) should be used and described in enough detail to reproduce. Report intercept and the coefficient of each variable.

#### 3.4 Risk Score Evaluation

After a risk score is constructed as detailed in Section 3.3 above, authors must outline the results of these models and all procedures utilized to validate the risk score. This includes whether the validation was performed on an internal sample or with external validation samples. All performance results should be described for both the development and validation samples either in the main text or supplement.

**Polygenic Risk Score Distribution.** The authors should provide a description, preferably with graphical representation, of the distribution of the risk score, as well as model fit measures. After individual genotypes are weighted by their effect sizes and summed across all sites (either genome-wide or select candidate sites), it is important to inspect the continuous distribution output directly from the risk model to check if assumptions are met, such as normality. Additionally, it is encouraged for authors to report the risk score distribution by case/control status, if applicable, as well as by any relevant variables such as sex, age, and/or race/ethnicity. If an integrated genetic risk model is utilized, authors should also describe the distribution of this risk score in addition to the genetics-only model to illustrate any differences that would contribute to changes in predictive ability.

**Risk Model Predictive Ability.** Authors should provide metrics of both overall performance (such as variance explained) and of estimated risk (relative or absolute). Once the continuous distribution of the risk score is calculated, it must be transformed into an individual's assessment of risk. This can take two forms: relative risk, which is in relation to a reference group, and absolute risk, which is in terms of their probability of an outcome. These two measures of risk require specific epidemiological and statistical considerations which must be addressed in the previous sections. If relative risk is utilized, authors should explicitly state the summary statistics used to estimate relative risk (e.g. hazard ratio (HR), odds ratio (OR) and/or regression coefficients ($\boldsymbol{\beta}$)). When stating this, authors should state any reference levels used (for instance: bottom third polygenic risk vs top third polygenic risk). See So & Sham, 2010 for details. [8] To this end, authors must fully describe the group definitions (both strata of interest and reference group) in terms of the items outlined in section *3.2 Study Populations*. This includes the risk score distribution stratified by risk groups and general population. Once this delineation has been determined, authors should provide enough detail should be included to enable readers to compute measures of risk score predictive ability such as sensitivity, specificity, positive predictive value (PPV), and negative predictive value (NPV). For absolute risk estimation, authors should describe the incidence/prevalence of the predicted outcome in the general population. If measures of risk are adjusted for other variables, these must be explicitly stated and described as per previous sections.

**Risk Model Discrimination.** Authors should describe and report metrics used to assess the discrimination of the risk score and whether any variables were included beyond the risk score in this analysis. This may include common metrics such as the area under the receiver operating characteristic curve (AUC/AUROC) or Precision-Recall (AUPRC), as well as the Concordance statistics (C-index) for survival models. This should also be presented as a visual or graphical display. While discrimination and calibration can be performed within the training set, it is most meaningful in an external validation.

**Risk Model Calibration.** Authors should describe and report metrics used to assess the calibration of the risk score and whether any variables were included beyond the risk score for this analysis. Metrics should be described and reported to test calibration for the constructed prediction model. This cannot be done for case-control validation cohorts, given the lack of a continuous outcome.

**Subgroup Analyses.** Often authors may test the performance of the developed PRS in a subgroup of the original study population. This offers perspective on the performance within subsets of participants as well as adjacent phenotypes. Authors should detail all subgroup analyses with the same level of detail described above for the main analysis. This includes all statistical methods used to estimate performance, as well as the risk score distribution, predictive ability, discrimination, and calibration. Subgroups should be defined and justified with consideration for downstream clinical utility.

#### 3.5 Limitations and Clinical Implications

Lastly, authors should provide a comprehensive and nuanced discussion of the broader context of their PRS, both in terms of target populations and possible downstream uses. It is important for authors to recognize potential limitations given the study population, availability of data, and/or statistical methods. By explicitly discussing these caveats, the PRS can be better contextualized, limiting the possibility of misuse.

**Risk Model Interpretation.** Authors should recap their study and summarize the risk model in terms of *how well* (prediction accuracy, discrimination, calibration) their risk model predicts *in whom* (study populations) and for *what* outcome (predicted end outcome). These definitions should be consistent with the concepts and motivations outlined in their introduction. Since many risk models are developed with an eye towards downstream clinical utility, it is important for authors to contextualize their findings in comparison to conventional clinical risk models. These conventional risk models may include demographic (age, sex), disease-specific risk factors, and/or family history of disease. Authors should describe the performance of both the PRS and clinical risk model (such as AUC), as well as the performance of the risk models combined if possible. Common comparisons that would compare the published PRS to standard-of-care in the field may be comparing the PRS to a family history of disease or known Mendelian variants with high penetrance. In all cases, the implications of these improvements should be outlined, such as reclassification metrics or the difference in proportion of risk/phenotypic variance explained.

**Limitations.** Authors must outline all limitations to their studies, including study design restrictions, ascertainment biases, the distribution of participant-level traits (ancestry, age, comorbidities), accuracy/specificity of phenotype data, and any statistical considerations. By discussing these limitations, authors will provide insight on the interpretation of the risk score, including both within the study population and to other target populations. Authors should include a discussion of any missing data or unknown reporting items from previous sections. Many of these limitations will likely pertain to the samples used in the development and validation of a PRS. While there should be no overlap between stages, independence is not always guaranteed due to large-scale consortiums combining different iterations of smaller studies. If there is any overlap this should be quantified and implications for interpretation should be outlined. The use of multi-center multi-study collaborations comes with some inherent caveats, including heterogeneity between studies in terms of study and participant-level traits. Authors must explain how this affects the confidence in prediction and how it influenced the methods utilized in the study, as well as any other caveats relevant to interpretation. For example, if data acquisition differs across the combined samples, explicitly state this. Additional common weaknesses in study design will heavily influence the estimation of relative and absolute risk. These may include biases in sample ascertainment due to recruitment method (e.g. convenience sampling) and recruitment setting (clinic vs. research vs. healthy populations). This is especially true if these methods impact disease prevalence metrics or the possibility of measuring secondary outcomes. Differences between study and target populations should be discussed, particularly in respect to these biases.

**Generalizability.** In addition to contextualizing the model within a risk model interpretation and recognizing its limitations, authors should discuss the generalizability beyond the study populations included in the development and validation of the PRS. Discussion should include additional populations and/or settings. If the risk score has been externally validated, authors should consider differences in performance with respect to ancestry, age, or other variables. Specific points should be made regarding the transportability of the PRS to other genetic ancestry groups, especially if the ancestry is not known or able to be disclosed within the study populations. Lastly, the discussion around generalizability should include whether previous findings were validated, such as previous GWAS or candidate gene studies. This content will further contextualize the PRS within the previously published body of work and the larger clinical field.

**Risk Model Intended Uses.** While every PRS may not have an intended clinical purpose or immediate clinical utility, authors should discuss if there is or what the downstream clinical purpose would be for their developed PRS. If there is an intended clinical purpose, authors must discuss the "clinical readiness" and the necessary next steps with respect to the interpretation, limitations, and generalizability of the model. Actionability should only be suggested and discussed if the study was set up appropriately with the relevant clinical population as the target of the specific PRS. The predictive ability of the model should be benchmarked against current standard of care or other published work (such as existing PRS) on predicting the clinical outcome of interest. This incremental value on top of established risk models for the disease assessed (e.g., Gail Model, 10-year CHD risk ACC/AHA pooled cohort equations). Additionally, authors should discuss meaningful risk reclassification, such as meeting specific treatment thresholds. If no clinical purpose is intended, authors should discuss the reasons prohibiting this use.

##### 3.6 Data Transparency and Availability.

Over the past several years, there has been a movement towards widespread data sharing with an emphasis on transparency and reproducibility. The ability of outside researchers to reproduce a PRS is vital for benchmarking against previously established risk assessments as well as future risk scores yet to be developed. Therefore, authors should supply sufficient information to calculate the PRS and/or risk model on external samples. For genetic variation, this would include information about the individual variants (e.g. rsID, chromosomal coordinates, effect allele, genome build, and effect weight with units), as well as the method used to combine these variant-level weights. This information should be shared in a publicly available database, such as the PGS Catalog [4] for findability and to promote re-use and comparison with other established scores. If other variables in addition to genetic variants are included in an integrated risk model, these effect sizes and data dictionary fields (including how variables are coded) should also be provided to ensure consistency.

Any risk scores that are intended for downstream clinical use should strive to meet stringent validation requirements as outlined in Roy et al (2018). [9]. Most notably this should include an evaluation of sensitivity, specificity, and positive predictive value to more easily facilitate clinical translation. Additional data helpful for external validation should be provided, including reference scores calculated on control or general population samples that are publicly available. Lastly, authors should clearly indicate how to find and access all of the data mentioned above.

#### 4. Manuscripts used to test framework for clarity and completeness

1. Belsky, D. W., M. R. Sears, R. J. Hancox, H. Harrington, R. Houts, T. E. Moffitt, K. Sugden, B. Williams, R. Poulton and A. Caspi (2013). "Polygenic risk and the development and course of asthma: an analysis of data from a four-decade longitudinal study." Lancet Respir Med 1(6): 453-461.
2. Choi, J., N. Song, S. Han, S. Chung, H. Sung, J. Y. Lee, S. Jung, S. K. Park, K. Y. Yoo, W. Han, J. W. Lee, D. Y. Noh, D. Kang and J. Y. Choi (2014). "The associations between immunity-related genes and breast cancer prognosis in Korean women." PLoS One 9(7): e103593.
3. Desikan, R. S., C. C. Fan, Y. Wang, A. J. Schork, H. J. Cabral, L. A. Cupples, W. K. Thompson, L. Besser, W. A. Kukull, D. Holland, C. H. Chen, J. B. Brewer, D. S. Karow, K. Kauppi, A. Witoelar, C. M. Karch, L. W. Bonham, J. S. Yokoyama, H. J. Rosen, B. L. Miller, W. P. Dillon, D. M. Wilson, C. P. Hess, M. Pericak-Vance, J. L. Haines, L. A. Farrer, R. Mayeux, J. Hardy, A. M. Goate, B. T. Hyman, G. D. Schellenberg, L. K. McEvoy, O. A. Andreassen and A. M. Dale (2017). "Genetic assessment of age-associated Alzheimer disease risk: Development and validation of a polygenic hazard score." PLoS Med 14(3): e1002258.
4. Ho-Le, T. P., J. R. Center, J. A. Eisman, H. T. Nguyen and T. V. Nguyen (2017). "Prediction of Bone Mineral Density and Fragility Fracture by Genetic Profiling." J Bone Miner Res 32(2): 285-293.
5. Holm, J., J. Li, H. Darabi, M. Eklund, M. Eriksson, K. Humphreys, P. Hall and K. Czene (2016). "Associations of Breast Cancer Risk Prediction Tools With Tumor Characteristics and Metastasis." J Clin Oncol 34(3): 251-258.
6. Inouye, M., G. Abraham, C. P. Nelson, A. M. Wood, M. J. Sweeting, F. Dudbridge, F. Y. Lai, S. Kaptoge, M. Brozynska, T. Wang, S. Ye, T. R. Webb, M. K. Rutter, I. Tzoulaki, R. S. Patel, R. J. F. Loos, B. Keavney, H. Hemingway, J. Thompson, H. Watkins, P. Deloukas, E. Di Angelantonio, A. S. Butterworth, J. Danesh, N. J. Samani and U. K. B. C. C. C. W. Group (2018). "Genomic Risk Prediction of Coronary

Artery Disease in 480,000 Adults: Implications for Primary Prevention." J Am Coll Cardiol 72(16): 1883-1893.

7. Kauppi, K., C. C. Fan, L. K. McEvoy, D. Holland, C. H. Tan, C. H. Chen, O. A. Andreassen, R. S. Desikan, A. M. Dale and I. Alzheimer's Disease Neuroimaging (2018). "Combining Polygenic Hazard Score With Volumetric MRI and Cognitive Measures Improves Prediction of Progression From Mild Cognitive Impairment to Alzheimer's Disease." Front Neurosci 12: 260.
8. Khera, A. V., C. A. Emdin, I. Drake, P. Natarajan, A. G. Bick, N. R. Cook, D. I. Chasman, U. Baber, R. Mehran, D. J. Rader, V. Fuster, E. Boerwinkle, O. Melander, M. Orho-Melander, P. M. Ridker and S. Kathiresan (2016). "Genetic Risk, Adherence to a Healthy Lifestyle, and Coronary Disease." N Engl J Med 375(24): 2349-2358.
9. Kuchenbaecker, K. B., L. McGuffog, D. Barrowdale, A. Lee, P. Soucy, J. Dennis, S. M. Dochek, M. Robson, A. B. Spurdle, S. J. Ramus, N. Mavaddat, M. B. Terry, S. L. Neuhausen, R. K. Schmutzler, J. Simard, P. D. P. Pharoah, K. Offit, F. J. Couch, G. Chenevix-Trench, D. F. Easton and A. C. Antoniou (2017). "Evaluation of Polygenic Risk Scores for Breast and Ovarian Cancer Risk Prediction in BRCA1 and BRCA2 Mutation Carriers." J Natl Cancer Inst 109(7).
10. Lacour, A., A. Espinosa, E. Louwersheimer, S. Heilmann, I. Hernandez, S. Wolfsgruber, V. Fernandez, H. Wagner, M. Rosende-Roca, A. Mauleon, S. Moreno-Grau, L. Vargas, Y. A. Pijnenburg, T. Koene, O. Rodriguez-Gomez, G. Ortega, S. Ruiz, H. Holstege, O. Sotolongo-Grau, J. Kornhuber, O. Peters, L. Frolich, M. Hull, E. Ruther, J. Wiltfang, M. Scherer, S. Riedel-Heller, M. Alegret, M. M. Nothen, P. Scheltens, M. Wagner, L. Tarraga, F. Jessen, M. Boada, W. Maier, W. M. van der Flier, T. Becker, A. Ramirez and A. Ruiz (2017). "Genome-wide significant risk factors for Alzheimer's disease: role in progression to dementia due to Alzheimer's disease among subjects with mild cognitive impairment." Mol Psychiatry 22(1): 153-160.
11. Lee, J., J. Choi, S. Chung, J. Park, J. E. Kim, H. Sung, W. Han, J. W. Lee, S. K. Park, M. K. Kim, S. H. Ahn, D. Y. Noh, K. Y. Yoo, D. Kang and J. Y. Choi (2017). "Genetic Predisposition of Polymorphisms in HMGB1-Related Genes to Breast Cancer Prognosis in Korean Women." J Breast Cancer 20(1): 27-34.
12. Li, H., B. Feng, A. Miron, X. Chen, J. Beesley, E. Bimeh, D. Barrowdale, E. M. John, M. B. Daly, I. L. Andrulis, S. S. Buys, P. Kraft, i. kConFab, H. Thorne, G. Chenevix-Trench, M. C. Southey, A. C. Antoniou, P. A. James, M. B. Terry, K. A. Phillips, J. L. Hopper, G. Mitchell and D. E. Goldgar (2017). "Breast cancer risk prediction using a polygenic risk score in the familial setting: a prospective study from the Breast Cancer Family Registry and kConFab." Genet Med 19(1): 30-35.
13. Maas, P., M. Barrdahl, A. D. Joshi, P. L. Auer, M. M. Gaudet, R. L. Milne, F. R. Schumacher, W. F. Anderson, D. Check, S. Chattopadhyay, L. Baglietto, C. D. Berg, S. J. Chanock, D. G. Cox, J. D. Figueroa, M. H. Gail, B. I. Graubard, C. A. Haiman, S. E. Hankinson, R. N. Hoover, C. Isaacs, L. N. Kolonel, L. Le Marchand, I. M. Lee, S. Lindstrom, K. Overvad, I. Romieu, M. J. Sanchez, M. C. Southey, D. O. Stram, R. Tumino, T. J. VanderWeele, W. C. Willett, S. Zhang, J. E. Buring, F. Canzian, S. M. Gapstur, B. E. Henderson, D. J. Hunter, G. G. Giles, R. L. Prentice, R. G. Ziegler, P. Kraft, M. Garcia-Closas and N. Chatterjee (2016). "Breast Cancer Risk From Modifiable and Nonmodifiable Risk Factors Among White Women in the United States." JAMA Oncol 2(10): 1295-1302.
14. Mavaddat, N., K. Michailidou, J. Dennis, M. Lush, L. Fachal, A. Lee, J. P. Tyrer, T. H. Chen, Q. Wang, M. K. Bolla, X. Yang, M. A. Adank, T. Ahearn, K. Aittomaki, J. Allen, I. L. Andrulis, H. Anton-Culver, N. N. Antonenkova, V. Arndt, K. J. Aronson, P. L. Auer, P. Auvinen, M. Barrdahl, L. E. Beane Freeman, M. W. Beckmann, S. Behrens, J. Benitez, M. Bermisheva, L. Bernstein, C. Blomqvist, N. V. Bogdanova, S. E. Bojesen, B. Bonanni, A. L. Borresen-Dale, H. Brauch, M. Bremer, H. Brenner, A. Brentnall, I. W. Brock, A. Brooks-Wilson, S. Y. Brucker, T. Bruning, B. Burwinkel, D. Campa, B. D. Carter, J. E. Castelao, S. J. Chanock, R. Chlebowski, H. Christiansen, C. L. Clarke, J. M. Collee, E. Cordina-Duverger, S. Cornelissen, F. J. Couch, A. Cox, S. S. Cross, K. Czene, M. B. Daly, P. Devilee, T. Dork, I. Dos-Santos-Silva, M. Dumont, L. Durcan, M. Dwek, D. M. Eccles, A. B. Ekici, A. H. Eliassen, C. Ellberg, C. Engel, M. Eriksson, D. G. Evans, P. A. Fasching, J. Figueroa, O. Fletcher, H. Flyger, A. Forsti, L. Fritschi, M. Gabrielson, M. Gago-Dominguez, S. M. Gapstur, J. A. Garcia-Saenz, M. M. Gaudet, V. Georgoulias, G. G. Giles, I. R. Gilyazova, G. Glendon, M. S. Goldberg, D. E. Goldgar, A. Gonzalez-Neira, G. I. Grenaker Alnaes, M. Grip, J. Gronwald, A. Grundy, P. Guenel, L. Haeberle, E. Hahnen, C. A. Haiman, N. Hakansson, U. Hamann, S. E. Hankinson, E. F. Harkness, S. N. Hart, W. He, A. Hein, J. Heyworth, P. Hillemanns, A. Hollestelle, M. J. Hooning, R. N. Hoover, J. L. Hopper, A. Howell, G. Huang, K. Humphreys, D. J. Hunter, M. Jakimovska, A. Jakubowska, W. Janni, E. M. John, N. Johnson, M. E. Jones, A. Jukkola-Vuorinen, A. Jung, R. Kaaks, K. Kaczmarek, V. Kataja, R. Keeman, M. J. Kerin, E.

Khusnutdinova, J. I. Kiiski, J. A. Knight, Y. D. Ko, V. M. Kosma, S. Koutros, V. N. Kristensen, U. Kruger, T. Kuhl, D. Lambrechts, L. Le Marchand, E. Lee, F. Lejbkowicz, J. Lilyquist, A. Lindblom, S. Lindstrom, J. Lissowska, W. Y. Lo, S. Loibl, J. Long, J. Lubinski, M. P. Lux, R. J. MacInnis, T. Maishman, E. Makalic, I. Maleva Kostovska, A. Mannermaa, S. Manoukian, S. Margolin, J. W. M. Martens, M. E. Martinez, D. Mavroudis, C. McLean, A. Meindl, U. Menon, P. Middha, N. Miller, F. Moreno, A. M. Mulligan, C. Mulot, V. M. Munoz-Garzon, S. L. Neuhausen, H. Nevanlinna, P. Neven, W. G. Newman, S. F. Nielsen, B. G. Nordestgaard, A. Norman, K. Offit, J. E. Olson, H. Olsson, N. Orr, V. S. Pankratz, T. W. Park-Simon, J. I. A. Perez, C. Perez-Barrios, P. Peterlongo, J. Peto, M. Pinchev, D. Plaseska-Karanfilska, E. C. Polley, R. Prentice, N. Presneau, D. Prokofyeva, K. Purrington, K. Pylkas, B. Rack, P. Radice, R. Rau-Murthy, G. Rennert, H. S. Rennert, V. Rhenius, M. Robson, A. Romero, K. J. Ruddy, M. Ruebner, E. Saloustros, D. P. Sandler, E. J. Sawyer, D. F. Schmidt, R. K. Schmutzler, A. Schneeweiss, M. J. Schoemaker, F. Schumacher, P. Schurmann, L. Schwentner, C. Scott, R. J. Scott, C. Seynaeve, M. Shah, M. E. Sherman, M. J. Shrubsole, X. O. Shu, S. Slager, A. Smeets, C. Sohn, P. Soucy, M. C. Southey, J. J. Spinelli, C. Stegmaier, J. Stone, A. J. Swerdlow, R. M. Tamimi, W. J. Tapper, J. A. Taylor, M. B. Terry, K. Thone, R. Tollenaar, I. Tomlinson, T. Truong, M. Tzardi, H. U. Ulmer, M. Untch, C. M. Vachon, E. M. van Veen, J. Vijai, C. R. Weinberg, C. Wendt, A. S. Whittemore, H. Wildiers, W. Willett, R. Winqvist, A. Wolk, X. R. Yang, D. Yannoukakos, Y. Zhang, W. Zheng, A. Ziogas, A. Investigators, A. I. kConFab, N. Collaborators, A. M. Dunning, D. J. Thompson, G. Chenevix-Trench, J. Chang-Claude, M. K. Schmidt, P. Hall, R. L. Milne, P. D. P. Pharoah, A. C. Antoniou, N. Chatterjee, P. Kraft, M. Garcia-Closas, J. Simard and D. F. Easton (2018). "Polygenic Risk Scores for Prediction of Breast Cancer and Breast Cancer Subtypes." Am J Hum Genet.

15. Mavaddat, N., P. D. Pharoah, K. Michailidou, J. Tyrer, M. N. Brook, M. K. Bolla, Q. Wang, J. Dennis, A. M. Dunning, M. Shah, R. Luben, J. Brown, S. E. Bojesen, B. G. Nordestgaard, S. F. Nielsen, H. Flyger, K. Czene, H. Darabi, M. Eriksson, J. Peto, I. Dos-Santos-Silva, F. Dudbridge, N. Johnson, M. K. Schmidt, A. Broeks, S. Verhoef, E. J. Rutgers, A. Swerdlow, A. Ashworth, N. Orr, M. J. Schoemaker, J. Figueroa, S. J. Chanock, L. Brinton, J. Lissowska, F. J. Couch, J. E. Olson, C. Vachon, V. S. Pankratz, D. Lambrechts, H. Wildiers, C. Van Ongeval, E. van Limbergen, V. Kristensen, G. Grenaker Alnaes, S. Nord, A. L. Borresen-Dale, H. Nevanlinna, T. A. Muranen, K. Aittomaki, C. Blomqvist, J. Chang-Claude, A. Rudolph, P. Seibold, D. Flesch-Janys, P. A. Fasching, L. Haeberle, A. B. Ekici, M. W. Beckmann, B. Burwinkel, F. Marme, A. Schneeweiss, C. Sohn, A. Trentham-Dietz, P. Newcomb, L. Titus, K. M. Egan, D. J. Hunter, S. Lindstrom, R. M. Tamimi, P. Kraft, N. Rahman, C. Turnbull, A. Renwick, S. Seal, J. Li, J. Liu, K. Humphreys, J. Benitez, M. Pilar Zamora, J. I. Arias Perez, P. Menendez, A. Jakubowska, J. Lubinski, K. Jaworska-Bieniek, K. Durda, N. V. Bogdanova, N. N. Antonenkova, T. Dork, H. Anton-Culver, S. L. Neuhausen, A. Ziogas, L. Bernstein, P. Devilee, R. A. Tollenaar, C. Seynaeve, C. J. van Asperen, A. Cox, S. S. Cross, M. W. Reed, E. Khusnutdinova, M. Bermisheva, D. Prokofyeva, Z. Takhirova, A. Meindl, R. K. Schmutzler, C. Sutter, R. Yang, P. Schurmann, M. Bremer, H. Christiansen, T. W. Park-Simon, P. Hillemanns, P. Guenel, T. Truong, F. Menegaux, M. Sanchez, P. Radice, P. Peterlongo, S. Manoukian, V. Pensotti, J. L. Hopper, H. Tsimiklis, C. Apicella, M. C. Southey, H. Brauch, T. Bruning, Y. D. Ko, A. J. Sigurdson, M. M. Doody, U. Hamann, D. Torres, H. U. Ulmer, A. Forsti, E. J. Sawyer, I. Tomlinson, M. J. Kerin, N. Miller, I. L. Andrulis, J. A. Knight, G. Glendon, A. Marie Mulligan, G. Chenevix-Trench, R. Balleine, G. G. Giles, R. L. Milne, C. McLean, A. Lindblom, S. Margolin, C. A. Haiman, B. E. Henderson, F. Schumacher, L. Le Marchand, U. Eilber, S. Wang-Gohrke, M. J. Hooning, A. Hollestelle, A. M. van den Ouweland, L. B. Koppert, J. Carpenter, C. Clarke, R. Scott, A. Mannermaa, V. Kataja, V. M. Kosma, J. M. Hartikainen, H. Brenner, V. Arndt, C. Stegmaier, A. Karina Dieffenbach, R. Winqvist, K. Pylkas, A. Jukkola-Vuorinen, M. Grip, K. Offit, J. Vijai, M. Robson, R. Rau-Murthy, M. Dwek, R. Swann, K. Annie Perkins, M. S. Goldberg, F. Labreche, M. Dumont, D. M. Eccles, W. J. Tapper, S. Rafiq, E. M. John, A. S. Whittemore, S. Slager, D. Yannoukakos, A. E. Toland, S. Yao, W. Zheng, S. L. Halverson, A. Gonzalez-Neira, G. Pita, M. Rosario Alonso, N. Alvarez, D. Herrero, D. C. Tessier, D. Vincent, F. Bacot, C. Luccarini, C. Baynes, S. Ahmed, M. Maranian, C. S. Healey, J. Simard, P. Hall, D. F. Easton and M. Garcia-Closas (2015). "Prediction of breast cancer risk based on profiling with common genetic variants." J Natl Cancer Inst 107(5).

16. Naslund-Koch, C., B. G. Nordestgaard and S. E. Bojesen (2017). "Common breast cancer risk alleles and risk assessment: a study on 35 441 individuals from the Danish general population." Ann Oncol 28(1): 175-181.

17. Natarajan, P., R. Young, N. O. Stitziel, S. Padmanabhan, U. Baber, R. Mehran, S. Sartori, V. Fuster, D. F. Reilly, A. Butterworth, D. J. Rader, I. Ford, N. Sattar and S. Kathiresan (2017). "Polygenic Risk Score

Identifies Subgroup With Higher Burden of Atherosclerosis and Greater Relative Benefit From Statin Therapy in the Primary Prevention Setting." Circulation 135(22): 2091-2101.

18. Paquette, M., M. Chong, S. Theriault, R. Dufour, G. Pare and A. Baass (2017). "Polygenic risk score predicts prevalence of cardiovascular disease in patients with familial hypercholesterolemia." J Clin Lipidol 11(3): 725-732 e725.
19. Paul, K. C., J. Schulz, J. M. Bronstein, C. M. Lill and B. R. Ritz (2018). "Association of Polygenic Risk Score With Cognitive Decline and Motor Progression in Parkinson Disease." JAMA Neurol 75(3): 360-366.
20. Ripatti, S., E. Tikkanen, M. Orho-Melander, A. S. Havulinna, K. Silander, A. Sharma, C. Guiducci, M. Perola, A. Jula, J. Sinisalo, M. L. Lokki, M. S. Nieminen, O. Melander, V. Salomaavea, L. Peltonen and S. Kathiresan (2010). "A multilocus genetic risk score for coronary heart disease: case-control and prospective cohort analyses." Lancet 376(9750): 1393-1400.
21. Rutten-Jacobs, L. C., S. C. Larsson, R. Malik, K. Rannikmae, M. consortium, C. International Stroke Genetics, C. L. Sudlow, M. Dichgans, H. S. Markus and M. Traylor (2018). "Genetic risk, incident stroke, and the benefits of adhering to a healthy lifestyle: cohort study of 306 473 UK Biobank participants." BMJ 363: k4168.
22. Seibert, T. M., C. C. Fan, Y. Wang, V. Zuber, R. Karunamuni, J. K. Parsons, R. A. Eeles, D. F. Easton, Z. Kote-Jarai, A. A. Al Olama, S. B. Garcia, K. Muir, H. Gronberg, F. Wiklund, M. Aly, J. Schleutker, C. Sipeky, T. L. Tammela, B. G. Nordestgaard, S. F. Nielsen, M. Weischer, R. Bisbjerg, M. A. Roder, P. Iversen, T. J. Key, R. C. Travis, D. E. Neal, J. L. Donovan, F. C. Hamdy, P. Pharoah, N. Pashayan, K. T. Khaw, C. Maier, W. Vogel, M. Luedeke, K. Herkommer, A. S. Kibel, C. Cybulski, D. Wokolorczyk, W. Kluzniak, L. Cannon-Albright, H. Brenner, K. Cuk, K. U. Saum, J. Y. Park, T. A. Sellers, C. Slavov, R. Kaneva, V. Mitev, J. Batra, J. A. Clements, A. Spurdle, M. R. Teixeira, P. Paulo, S. Maia, H. Pandha, A. Michael, A. Kierzek, D. S. Karow, I. G. Mills, O. A. Andreassen, A. M. Dale and P. Consortium* (2018). "Polygenic hazard score to guide screening for aggressive prostate cancer: development and validation in large scale cohorts." BMJ 360: j5757.
23. Song, N., K. Kim, A. Shin, J. W. Park, H. J. Chang, J. Shi, Q. Cai, D. Y. Kim, W. Zheng and J. H. Oh (2018). "Colorectal cancer susceptibility loci and influence on survival." Genes Chromosomes Cancer 57(12): 630-637.
24. Tada, H., O. Melander, J. Z. Louie, J. J. Catanese, C. M. Rowland, J. J. Devlin, S. Kathiresan and D. Shiffman (2016). "Risk prediction by genetic risk scores for coronary heart disease is independent of self-reported family history." Eur Heart J 37(6): 561-567.
25. Talmud, P. J., J. A. Cooper, J. Palmen, R. Lovering, F. Drenos, A. D. Hingorani and S. E. Humphries (2008). "Chromosome 9p21.3 coronary heart disease locus genotype and prospective risk of CHD in healthy middle-aged men." Clin Chem 54(3): 467-474.
26. Tan, C. H., B. T. Hyman, J. J. X. Tan, C. P. Hess, W. P. Dillon, G. D. Schellenberg, L. M. Besser, W. A. Kukull, K. Kauppi, L. K. McEvoy, O. A. Andreassen, A. M. Dale, C. C. Fan and R. S. Desikan (2017). "Polygenic hazard scores in preclinical Alzheimer disease." Ann Neurol 82(3): 484-488.
27. Ten Broeke, S. W., F. A. Elsayed, L. Pagan, M. J. W. Olderode-Berends, E. G. Garcia, H. J. P. Gille, L. P. van Hest, T. G. W. Letteboer, L. E. van der Kolk, A. R. Mensenkamp, T. A. van Os, L. Spruijt, B. J. W. Redeker, M. Suerink, Y. J. Vos, A. Wagner, J. T. Wijnen, E. W. Steyerberg, C. M. J. Tops, T. van Wezel and M. Nielsen (2018). "SNP association study in PMS2-associated Lynch syndrome." Fam Cancer 17(4): 507-515.
28. Vaara, S., E. Tikkanen, O. Parkkonen, M. L. Lokki, S. Ripatti, M. Perola, M. S. Nieminen and J. Sinisalo (2016). "Genetic Risk Scores Predict Recurrence of Acute Coronary Syndrome." Circ Cardiovasc Genet 9(2): 172-178.
29. Vrshek-Schallhorn, S., C. B. Stroud, S. Mineka, R. E. Zinbarg, E. K. Adam, E. E. Redei, C. Hammen and M. G. Craske (2015). "Additive genetic risk from five serotonin system polymorphisms interacts with interpersonal stress to predict depression." J Abnorm Psychol 124(4): 776-790.
30. Wimberley, T., C. Gasse, S. M. Meier, E. Agerbo, J. H. MacCabe and H. T. Horsdal (2017). "Polygenic Risk Score for Schizophrenia and Treatment-Resistant Schizophrenia." Schizophr Bull 43(5): 1064-1069.
